# A curated reference dataset and deep learning model for multi-lead electrocardiographic interval measurements in UK Biobank

**DOI:** 10.64898/2026.06.26.26356665

**Authors:** Thomas Kaplan, Julia Ramírez, William J Young, Mihir M Sanghvi, Josseline Madrid, Hafiz Naderi, Ravi Shah, Ana Minchole, Michele Orini, Andrew Tinker, Pier D Lambiase, Patricia B Munroe, Stefan van Duijvenboden

## Abstract

Electrocardiographic (ECG) interval measurements underpin clinical decision-making and large-scale cardiovascular research, yet existing automated methods are often developed using small, heterogeneous datasets with limited expert annotation and uncertain generalisability to population cohorts. We developed a deep learning framework for automated PR, QRS, and QT interval estimation and established a large expert-curated reference dataset using UK Biobank (UKB) ECGs. The reference dataset comprises 11,330 lead-level annotations from 12-lead ECGs in 1,030 randomly selected UKB participants, generated using a standardised annotation protocol with independent expert review. A 1D convolutional neural network was trained to segment ECG waveforms and derive PR, QRS, and QT intervals. Performance was evaluated against expert annotations, UKB CardioSoft measurements, an open-source signal-processing toolbox, and a wavelet-based delineation method. Clinical validity was evaluated through associations with incident atrial fibrillation and major adverse cardiovascular events (MACE). Inter-observer agreement was high (ICC 0.81–0.97). In a held-out test set, the deep learning model achieved mean absolute errors of 7.7 ms (PR), 7.5 ms (QRS), and 4.9 ms (QT), outperforming all comparator methods, with minimal bias relative to expert annotations. Review of distributional outliers confirmed >80% validity for most interval measurements. Among 46,749 participants with follow-up (median 4 years), prolonged QTc derived by the deep learning model showed stronger associations with incident MACE (hazard ratio 2.9, 95% CI 2.1–4.0) than wavelet-based measurements (1.7, 1.4–2.0) or CardioSoft (1.1, 0.9–1.4). This expert-curated reference dataset and validated deep learning framework provide a scalable foundation for reproducible ECG phenotyping in UKB and beyond.

## Introduction

Understanding cardiac electrical behaviour is fundamental to cardiovascular medicine and population health research. The electrocardiogram (ECG) provides a simple, non-invasive means of capturing cardiac activation and recovery, and has shown to be a powerful tool in large-scale genetic and epidemiological studies. Among the most informative clinical ECG-derived traits are the PR, QRS, and QT intervals, which reflect atrioventricular conduction and ventricular depolarisation and repolarisation, respectively. Abnormal intervals have been associated with arrhythmic risk, heart failure, and other adverse cardiovascular events, providing key insights into pathophysiological pathways and potential targets for prevention[1–5]. Consequently, accurate ECG delineation is fundamental in enabling a wide range of downstream clinical and population-based research studies.The UK Biobank (UKB) has recently completed 12-lead ECG acquisition in 100,000 participants, creating a major resource for population scale investigation of ECG traits linked to extensive ‘omics and phenotypic data[6]. While automatic interval measurements from the acquisition system (Cardiosoft) are available, their accuracy and reproducibility have not been systematically validated. Similar uncertainty applies to accessible alternatives, including open-source toolboxes such as *NeuroKit*[7], which have not been specifically calibrated or benchmarked for UKB data. Separately, recent deep learning delineation models have achieved near expert performance in large clinical datasets[8–10], but their translation to UKB is not straightforward because these models were trained and validated on data acquired using different hardware, preprocessing pipelines, populations, and annotation conventions. Furthermore, the corresponding reproducible implementations are often not publicly available. Without specific validation and a transparent delineation pipeline, confidence in ECG interval estimation will remain limited, constraining downstream epidemiological and omics analyses. Importantly, these challenges are not unique to UKB, and methods validated in UKB may enable scalable ECG phenotyping in other population cohorts with minimal fine-tuning.

In this study, we address these limitations with two key contributions. First, we constructed a large reference dataset with curated PR, QRS, and QT measurements derived from 11,330 signal-averaged ECG waveforms across independent leads (I, II, V1–V6) and computed orthogonal leads (X, Y, and Z). These waveforms were obtained from 10 second resting 12-lead ECGs recorded in 1,030 randomly selected UKB participants with high quality, artefact free signals. Second, we developed a deep learning waveform segmentation model to enable accurate and automated estimation of PR, QRS, and QT onsets, offsets, and intervals. We will make this algorithm and curated data available through UKB and open-access repositories, establishing the first openly available, validated framework for ECG interval measurement in UKB. This resource provides a standardised foundation for reproducible ECG trait extraction and advances large-scale population-based cardiovascular research.

## Methods

### Study population

The UKB is a large prospective cohort of approximately 500,000 participants recruited between 2006 and 2010 to investigate genetic, environmental, and lifestyle determinants of health and disease. An imaging extension of the study, launched in 2014 with the aim of scanning 100,000 participants, including repeat assessments in up to 60 000 participants, acquired standard 12-lead resting ECGs using a standardised protocol. (GE Cardiac Acquisition Module CAM-14; https://biobank.ctsu.ox.ac.uk/ukb/ukb/docs/12lead_ecg_explan_doc.pdf). This dataset comprises raw ECG waveforms sampled at 500 Hz, together with interval measurements automatically derived by the interpretation software (GE CardioSoft Version 6 system; https://biobank.ctsu.ox.ac.uk/crystal/ukb/docs/CardiosoftFormatECG.pdf).

### ECG preprocessing

For each participant, we analysed 10-second raw ECG recordings to construct signal-averaged median waveforms. ECGs were first zero-phase band-pass filtered between 1 and 45 Hz using a fourth-order Butterworth filter to remove baseline wander and high-frequency noise. R-waves were then detected in lead I for each normal heartbeat using a previously validated algorithm[11]. Individual beats were aligned to the detected R wave and combined within each lead to derive a median P, QRS, and T waveform for subsequent analyses. In addition to the recorded 12 leads, orthogonal X, Y, and Z vectorcardiographic leads were reconstructed using the Kors regression matrix[12]. To minimise redundancy and collinearity among highly correlated derived leads, analyses in this work were restricted to the independent limb and precordial leads I, II, and V1 to V6, together with the orthogonal X, Y, and Z leads.

### Selection of reference ECGs

From 73,149 UK Biobank ECG recordings available at the time of analysis, we applied stringent quality-control procedures to derive a high-quality subset of rhythm-stable median beats for expert annotation, providing reliable training data for segmentation modelling. First, duplicate recordings were removed (n = 4,454). We then selected median waveforms derived from rhythm-stable ECG recordings, defined as those containing ≥5 analysable beats, a mean RR interval between 500–1500ms, limited RR variability (max–min RR <200ms), and adequate signal quality across all considered leads (QRS amplitude >0.1 mV; SNR >0 dB). After exclusions, 52,746 ECGs remained (Figure 1). From this subset, 1,030 recordings were randomly selected to create the curated reference dataset. Of these, 1,000 ECGs were distributed across eight reviewers (125 each), and 30 were reviewed by all reviewers to assess inter observer variability, yielding 11,330 annotated lead level recordings. All reviewers were trained ECG annotators.

**Figure 1:**
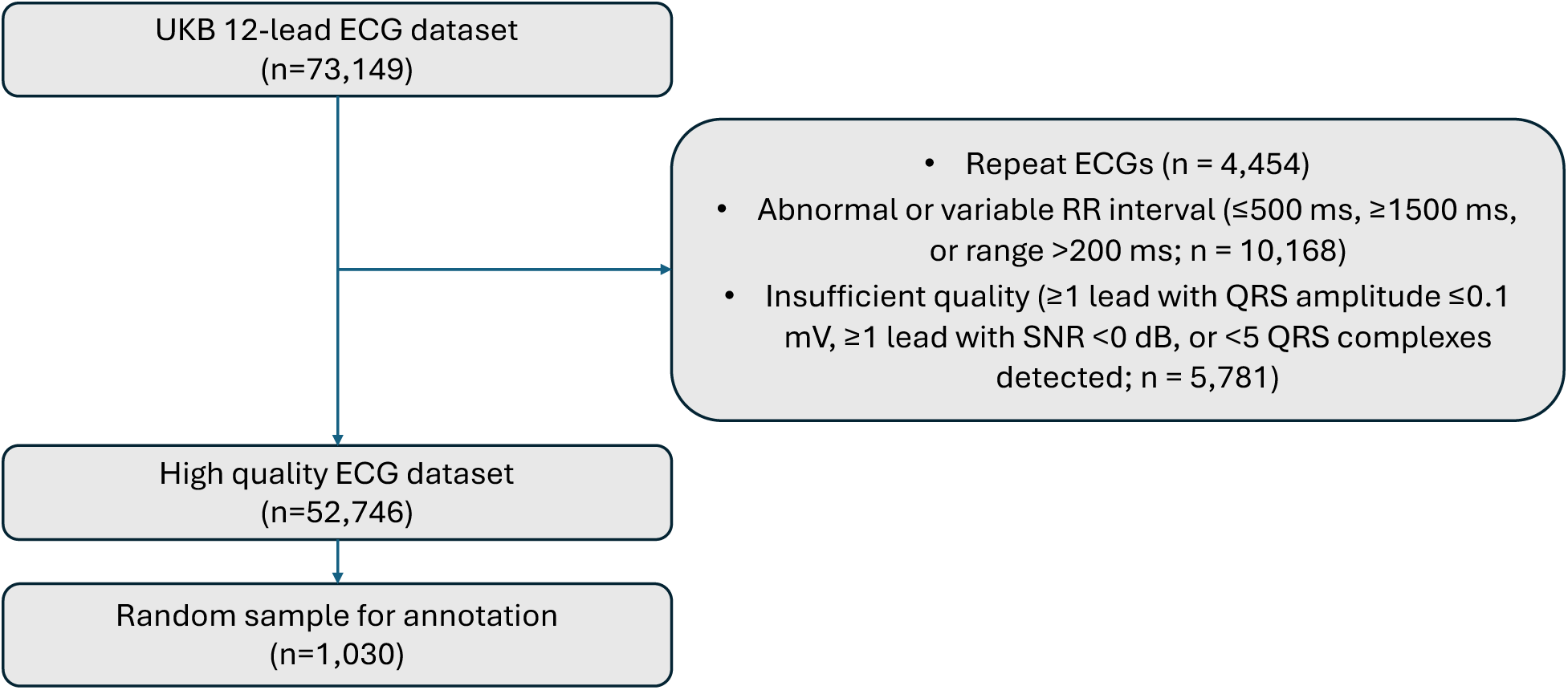
Selection of UK Biobank 12-lead ECG recordings for manual annotation. After exclusions, 52,746 quality-controlled ECGs were retained, from which 1,030 were randomly selected for annotation; 1,000 were distributed evenly among eight reviewers (n = 125), and 30 were reviewed by all to assess inter-observer variability.

### Expert annotation protocol

ECGs were annotated independently by the reviewers using a standardised protocol and a dedicated graphical user interface created using Python (v3.10) (Supplemental Figure 1). For each recording, the onset and offset of each P-wave, QRS complex, and T-wave were annotated. Annotations for the P-waves and QRS complexes were made once per recording, as PR interval and QRS duration exhibit minimal inter lead variability. In contrast, T-wave annotations were performed independently across the selected leads (I, II, V1–V6, X, Y, and Z), as T-wave morphology and repolarisation timing can differ substantially between leads, as observed clinically with QT dispersion[13]. The end of the T wave was defined using the tangent method, whereby a tangent is drawn to the steepest downslope of the last limb of the T wave and its intersection with the isoelectric baseline as the endpoint, as recommended in established guidelines[14]. When the onset or offset of a waveform could not be clearly identified (e.g. low amplitude, noise, or complex morphology) annotators flagged the segment as uncertain, and such markers were left unannotated for downstream analyses. Prior to annotating the 125 assigned ECG recordings, reviewers conducted a calibration exercise involving joint review and consensus discussion to harmonise annotation criteria and ensure consistency across observers (Supplemental Methods).

### Deep learning segmentation model

We developed a one-dimensional convolutional neural network (1D CNN) to segment ECG waveforms and derive the onset and offset of the P wave, QRS complex, and T wave. The network architecture (Supplemental Figure 2) consisted of multiple convolutional layers operating at progressively increasing temporal scales, followed by dilated convolutions to capture mid– and long-range dependencies, and a final dense layer with softmax activation to output class probabilities per time step. The model was trained on the expert-annotated reference dataset; for the 30 ECGs reviewed by all annotators, median onset and offset timings across reviewers were used as consensus labels. Data were randomly partitioned into training and test sets without stratification by annotating expert with 70% of the ECGs used for model training, corresponding to N=7931 lead recordings, and the remainder used for comparison of manual and automated annotations. A single standardised median ECG beat (standard deviation 1, mean 0) was used as input, while the corresponding time-step labels served as supervision targets.

Model training was performed in Python (v3.10) using TensorFlow (v2.16.2) and Keras (v3.6.0) on the Queen Mary University of London Apocrita high-performance computing facility. Post-processing included several steps: interval probabilities outside valid sample windows were zeroed, the most likely interval class per sample was assigned, and the resulting class sequence was smoothed with a median filter to extract contiguous waveform boundaries and calculate PR, QRS, and QT intervals. Detailed information on model architecture, hyperparameter optimisation, training procedure, and post-processing algorithms is provided in the Supplementary Methods.

### Benchmarking and comparative methods

Model performance was benchmarked against the expert-annotated reference dataset and compared with three existing approaches: (i) measurements provided by the ECG interpretation software (GE CardioSoft version 6 system) used by and available through the UKB, (ii) automated delineation using the open-source Python package NeuroKit (v0.2.10)[7], and (iii) a wavelet-based segmentation algorithm previously developed by Martínez et al.[15] at the University of Zaragoza, here referred to as the “wavelet-based” method (as implemented in a recent study[16]).

CardioSoft-derived intervals were obtained directly from the XML output files available in the UKB resource (https://biobank.ndph.ox.ac.uk/ukb/ukb/docs/CardiosoftFormatECG.pdf). NeuroKit was applied using its standard ECG delineation pipeline, which includes signal pre-processing, QRS detection, and phase segmentation. The wavelet-based method was implemented in MATLAB (R2022b, The MathWorks Inc.) following the original publication, with interval-specific thresholds applied for PR, QRS, and QT segmentation. Full implementation details and parameter settings for all methods are provided in the Supplementary Methods.

### Statistical analyses

Agreement between automated and manual ECG annotations was assessed using intra-class correlation coefficients (ICC) based on a two-way mixed-effects consistency model (ICC[3,1])[17], implemented in Pingouin (v0.5.5). Pearson correlation coefficients were computed using SciPy (v1.15.1), and differences between correlations were tested using Fisher’s Z-transformation. Agreement was further evaluated using ordinary least squares (OLS) regression (statsmodels, v0.14.4). Further details are provided in Supplementary Methods.

To evaluate clinical validity, we examined established associations between PR interval and incident atrial fibrillation, and between QRS duration and QT interval and major adverse cardiovascular events (MACE) using the hospital episode statistics available in UKB with censoring applied at 31 October 2022 (definitions provided in Supplemental Tables 1 and 2). As exposures we used first degree atrioventricular block, prolonged QRS, and prolonged corrected QT were defined according to contemporary guideline thresholds: PR > 200 ms[18], QRS > 120 ms[19], and QTc > 460 ms in women and > 440 ms in men[20]. Survival analyses were performed in R (v4.5.1) using RStudio (2025). Cox proportional hazards regression models were fitted using the survival (v3.8-3) and survminer (v0.5.0) packages. Participants were excluded if their censoring date preceded the date of the ECG recording or if they had a prior history of cardiovascular disease (Definitions are provided in Supplementary Table 3). The orthogonal leads (X, Y, Z) were excluded for comparability as they were not made available for the wavelet-based method, and analyses were restricted to participants with at least one successfully annotated lead (n = 57,552). Both univariate and multivariate models adjusted for age and sex were evaluated. In addition to outcome associations, we also evaluated the physiological validity of PR and QT intervals by visually inspecting their relationships with heart rate, age, and sex, given their well-established rate and demographic dependencies.

### Outlier review

To assess the validity of automatically derived interval measurements at extreme values, we conducted a targeted review of outlier ECGs. For each interval type (PR, QRS, and QT), recordings within the top and bottom 1% of the respective distributions were identified on a lead-independent basis. These ECGs were reviewed by five of the eight expert annotators following a standardised protocol, (the clinical practising cardiologists in the team. Reviewers were provided with a graphical interface similar to that used during annotation, displaying all available leads (I, II, V1–V6, X, Y, and Z), but with the lead containing the outlier measurement highlighted. Each reviewer was assigned one or two interval types (PR, QRS, and/or QT) and classified outliers as valid, unclear, or artefactual (indicative of waveform corruption). We then quantified the proportion in each interval category.

## Results

In the curated reference ECG dataset, the mean age of participants was 65·5 years (SD 7·7), and 566 (55·0%) of the 1,030 participants were women. Sex-stratified PR, QRS, and QT interval measurements are summarised in Table 1. PR interval and QRS duration, annotated once per recording on a lead independent basis, were measurable in 1,015 (96.7%) and 1,028 (99.8%) of the 1,030 ECG recordings, respectively. Among women, the mean PR interval was 161.9 ± 24.4ms and the mean QRS duration was 93.9 ± 12.3ms. Among men, the corresponding values were 177.1 ± 30.3ms and 102.1 ± 17.2 ms. QT interval measurements were available in more than 95% of recordings for each lead, except leads V1 (91.0%) and Z (94.0%). The mean QT interval (across all leads) was 397.1 ± 30.8ms in women and 394.1 ± 33.8ms in men, corresponding to mean corrected QT intervals (QTc) of 403.4 ± 23.1ms and 392.2 ± 25.1ms, respectively.

**Table 1:** ECG intervals in the curated data set (N=1,030 ECG).

|  | **Women (n=566, 55.0%)** | **Avail. (%)** | **Men (464, 45.0%)** | **Avail. (%)** |
| --- | --- | --- | --- | --- |
| **PR interval (ms)** | 161.9 ± 24.4 | 99.3 | 177.1 ± 30.3 | 99.3 |
| **QRS duration (ms)** | 93.9 ± 12.3 | 99.8 | 102.1 ± 17.2 | 99.8 |
| **QT interval (ms)** |  |  |  |  |
| I | 395.6 ± 30.0 | 97.9 | 391.6 ± 32.8 | 97.0 |
| II | 398.6 ± 29.5 | 98.8 | 394.2 ± 32.8 | 98.3 |
| V1 | 392.0 ± 33.3 | 91.5 | 385.5 ± 35.5 | 90.3 |
| V2 | 390.7 ± 30.5 | 95.1 | 389.3 ± 32.7 | 97.0 |
| V3 | 400.4 ± 29.5 | 96.8 | 395.6 ± 33.9 | 99.6 |
| V4 | 401.7 ± 29.8 | 97.2 | 398.9 ± 32.9 | 99.6 |
| V5 | 401.2 ± 30.5 | 97.9 | 398.9 ± 32.9 | 98.9 |
| V6 | 400.4 ± 30.6 | 97.9 | 398.3 ± 33.2 | 98.3 |
| X | 396.9 ± 30.8 | 95.8 | 396.1 ± 33.2 | 97.6 |
| Y | 399.7 ± 31.1 | 98.1 | 397.9 ± 34.2 | 98.7 |
| Z | 390.0 ± 30.5 | 94.5 | 387.7 ± 34.8 | 93.3 |

### Inter-observer variability

Among the 30 ECG recordings reviewed independently by all annotators, good agreement was observed across all ECG interval measurements, with ICCs of 0.87 (95% CI 0.71–0.97) for PR interval, 0.81 (0.65–0.90) for QRS duration, and 0.97 (0.95–0.98) for QT interval. For PR interval and QRS duration, which were reviewed once per recording on a lead independent basis, the mean absolute pairwise difference was 7.1ms and 6.7ms, respectively. For QT interval, which was reviewed on a lead specific basis (11 leads per ECG recording), the mean difference ranged between 4.2ms (lead I) and 6.5ms (lead V1).

### Automatic and manual annotation agreement

An example ECG annotation generated by the deep-learning method is shown in Figure 2, displaying the ECG waveform with overlaid probability maps for P, QRS, and T waves, alongside the final annotations derived from the highest-probability predictions at each timepoint. Annotations for the N=309 hold-out ECGs generated by our deep-learning method, wavelet-based method, and UKB Cardiosoft all demonstrated strong agreement, correlation and linear fits (Supplemental Tables 4 and 5). Mean absolute errors (MAE) for intervals averaged across leads were superior in the case of the deep-learning method, at MAE=7.7ms [SD=6.5] for PR, MAE=7.5ms [SD=6.5] for QRS duration and MAE=4.9ms [SD=4.1] for QT interval (Table 2). Automated annotation was most performant for QT interval, followed by PR interval and QRS duration. For averaged intervals across leads (Supplemental Table 4), correlations with manual annotations were stronger for our deep-learning method than for UKB-CardioSoft for both PR (z=-2.46, p=0.014) and QT intervals (z=-6.23, p<0·001), and stronger than the wavelet-based method for PR interval (z=-3.12, p=0.002). Correlations for QT interval did not differ significantly from the wavelet-based method (z=-1.89, p=0.059), and correlations for QRS duration were similar across methods (CardioSoft: z=-1.04, p=0.297; wavelet-based method: z=-1.89, p=0.059). Whilst NeuroKit demonstrated poor correlation between lead-averaged annotations and that of manual annotations, all metrics saw improvements in lead-specific QT annotations.

**Figure 2:**
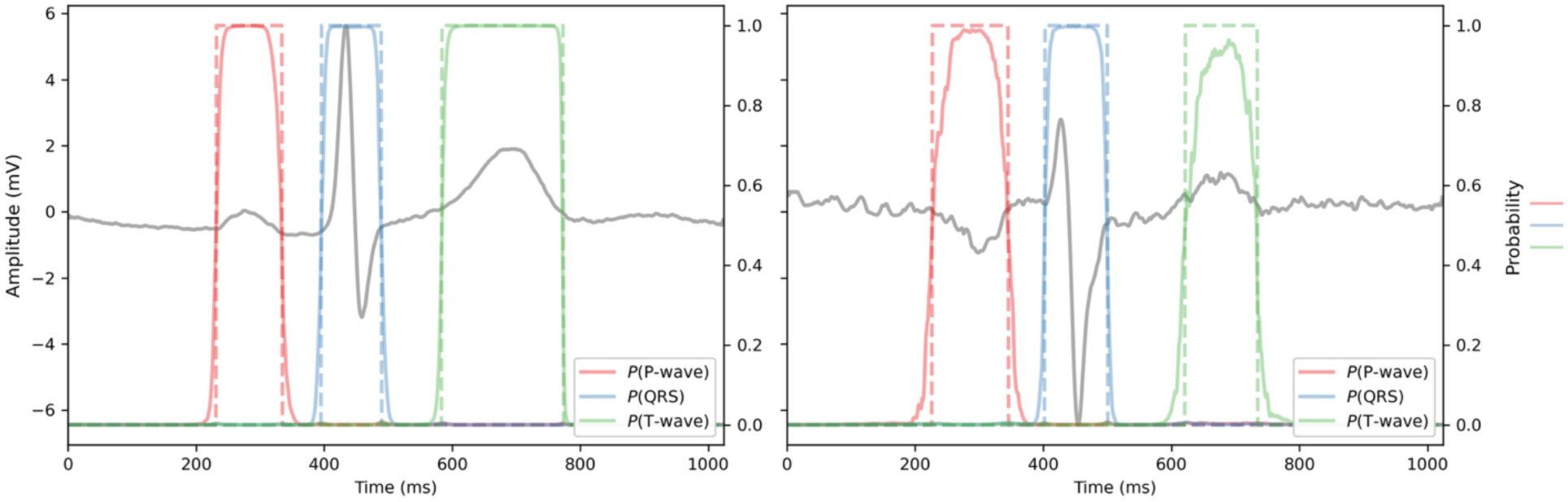
Examples of ECG annotation with overlaid underlying probability maps, alongside the argmax used for interval derivation. The right example demonstrates how the probability sequence degrades due to signal ambiguity (e.g. noise), such that the probability map (solid line) exhibits noise and diverging from the argument of the maximum (dashed line).

**Table 2:** Mean absolute errors (MAE) of lead-averaged intervals with respect to the curated dataset (N=309).

| ***Interval*** | ***Method*** | **MAE (ms)** | **SD** |
| --- | --- | --- | --- |
| *PR* | *Deep-learning (present work)* | 7.69 | 6.53 |
|  | *Wavelet-based* | 14.10 | 10.69 |
|  | *UKB CardioSoft* | 9.41 | 8.21 |
| *QRSduration* | *Deep-learning (present work)* | 7.53 | 6.48 |
|  | *Wavelet-based* | 12.17 | 9.18 |
|  | *UKB CardioSoft* | 11.61 | 8.77 |
| *QT* | *Deep-learning (present work)* | 4.91 | 4.08 |
|  | *Wavelet-based* | 11.65 | 9.32 |
|  | *UKB CardioSoft* | 23.76 | 9.73 |

Corresponding Bland-Altman analyses (Figure 3 and Supplemental Figure 3) further demonstrated the precision of automated annotations generated in the present work, highlighting marginal mean differences between the annotations. For the deep-learning method, the mean differences with the manual annotation were 0.6ms (95% CI: –19.2 – 20.3), –0.7ms (–20.6 – 18.7), and –1.5ms (–13.7 – 10.6) for PR interval, QRS duration, and QT interval, respectively. In comparison, UKB CardioSoft results demonstrated wider limits of agreement and a systematic bias for QRS duration (–9.3 ms) and QT interval (23.5 ms; Figure 3). A similar directional bias was also observed for the Wavelet-based method (Figure 3). Note that the diagonal series of markers have two underlying causes: (1) the expert annotation interface enforced a single PR and QRS duration interval across leads, but it was possible for lead-specific automatic annotations to differ; (2) at extremes, method-specific interval thresholds are applied (see Methods).

**Figure 3:**
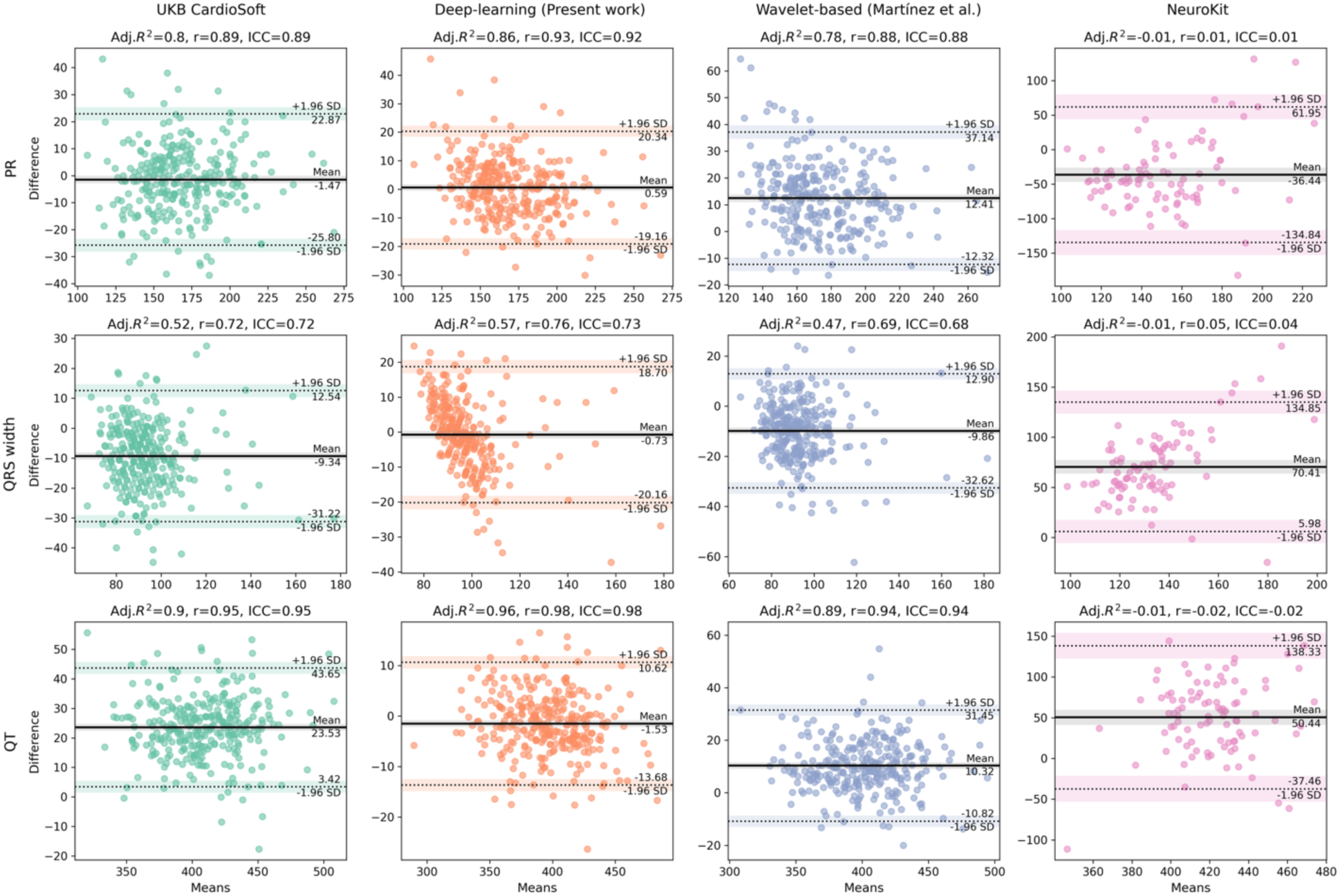
Bland-Altman analysis for pairwise comparison of automated annotations (median interval across leads) and expert (manual) annotations. Corresponding agreement statistics are reported in **Error! Reference source not found.**.

As a rough indication of relative runtime performance, using 4 vCPUs each with 16GB RAM, the average signal annotation time was 0.16 seconds for the deep-learning method (present work); 0.24 seconds for the wavelet-based method; and 0.54 seconds for NeuroKit.

### Automatic annotation outlier assessment

On average, each expert reviewer assessed approximately 4,750 automatically annotated waveforms, and most outlier categories demonstrated over 80% validity (Supplementary Figure 4). The one notable exception was that of short QT annotations, where just ∼60% were deemed valid, and with negligible corrupt or ambiguous annotations. As shown in Supplemental Figure 5, nearly all automated QT interval annotations <250ms were classified as invalid, whereas measurements exceeding this threshold were increasingly likely to be considered valid and within physiological ranges. A similar relationship between annotation validity and interval length is shown in most outlier categories (Supplemental Figure 5).

### Validation of annotation associations

Among 63,790 participants with valid interval measurements in at least one lead across all annotation approaches, application of predefined exclusion criteria (prior cardiovascular disease, no follow-up beyond baseline, heart rate <40 or >120 bpm) yielded a cohort of 46,749 participants with follow-up data. The three methods were able to annotate an interval in at least one lead for the majority of participants (N=[46,715–46,749], 99-100% of this sub-cohort), with the exception of CardioSoft, whereby 3.8% (N=1757) individuals did not have an annotated PR interval. During a median follow-up of 3.97 years (IQR 2.58), there were 811 AF events and 725 MACE events. Overall, the direction of associations was broadly consistent across the deep learning, wavelet-based, and UKB Cardiosoft algorithms; however, ECG intervals derived using the deep learning and wavelet-based approaches generally showed stronger and more consistent associations with cardiovascular outcomes than UKB Cardiosoft annotations in age– and sex-adjusted Cox proportional hazards models (Figure 4; Supplemental Tables 6 and 7). In particular, deep learning-derived measurements of prolonged QTc interval yielded substantially larger effect estimates for incident MACE (HR 2.9, 95% CI 2.1–4.0) than the wavelet-based method (HR 1.7, 95% CI 1.4–2.0), whereas no association was observed for corresponding UKB Cardiosoft-derived measurements (1.1, 0.9–1.4; Figure 4; Supplemental Table 7).We also observed that PR and QT interval annotations in the present work do conform to the expected relationship with heart rate (Supplemental Figures 6-8). Similar relationships were observed with the wavelet-based method and UKB CardioSoft (Supplemental Figure 8), again with the systematic bias observed earlier in the earlier Bland–Altman comparisons against expert annotations (Figure 3).

**Figure 4:**
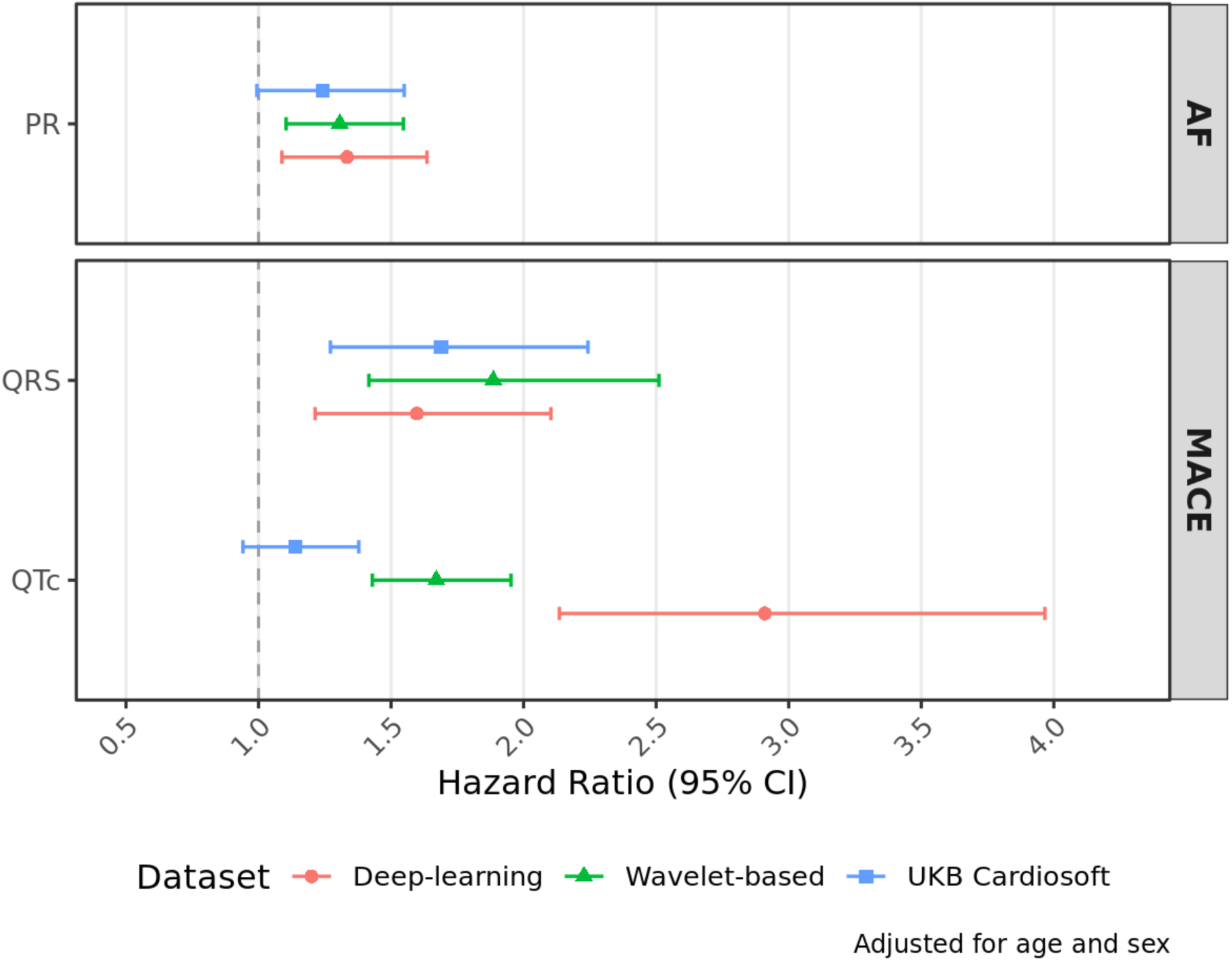
Outcome associations for multivariate Cox proportional hazards regression models for atrial fibrillation (AF) and Major Adverse Cardiovascular Events in medicine (MACE), adjusted for age and sex. Error bars correspond to 95% confidence intervals.

## Discussion

By integrating expert-led annotation, systematic benchmarking against established approaches, and downstream validation using clinical outcomes, the present work addresses a key methodological gap that has constrained the reproducibility and interpretability of ECG-derived phenotypes in large population studies, including UKB. Reliable large-scale ECG delineation is important because interval-derived phenotypes underpin epidemiological, genetic, and drug discovery analyses across population cohorts[3,21–24]. Our findings demonstrate that deep-learning–based delineation can achieve high agreement with expert annotation, outperform commonly used automated pipelines, and yield interval estimates that retain expected physiological relationships and clinically meaningful associations. The curated dataset showed high measurement completeness and strong inter-observer consistency, supporting the robustness of the expert annotations as a reference standard. Together, these resources provide a scalable framework for reproducible ECG phenotyping in UKB and beyond, facilitating more reliable investigation of cardiac electrophysiology across large population cohorts.

Our deep learning framework demonstrated closer agreement with expert annotation than both the wavelet-based approach and the interval measurements currently provided through UKB (CardioSoft), particularly for PR and QT interval estimation. Although wavelet-based methods have historically underpinned many automated ECG delineation pipelines[15,25], our findings support growing evidence that deep learning approaches may offer greater robustness across the waveform variability encountered in large population datasets[10]. Reduced systematic bias and stronger preservation of expected physiological and clinical associations further support the utility of deep learning–based delineation for large-scale ECG phenotyping. This was particularly evident for QTc interval, where stronger associations with MACE for the deep-learning method may reflect reduced threshold misclassification compared with alternative approaches. As preprocessing pipelines and waveform selection criteria were broadly similar across methods, the observed differences are unlikely to be explained by preprocessing alone. Some divergence would nevertheless be expected because the deep learning model was trained directly on UKB-derived expert annotations, whereas the wavelet-based method was developed using different datasets with different delineation conventions and lead configurations, including PhysioNet[15,26].

Recent studies have shown that convolutional segmentation models can achieve strong performance on benchmark datasets. For example, Chen et al. and Joung et al, reported that a 1D-UNet-based approach, enhanced by single-beat segmentation and morphology-informed post-processing, substantially improved ECG delineation accuracy for P, QRS, and T boundaries[10,27]. However, in contrast to our work, where mean absolute errors were below 10ms, these studies often evaluate delineation performance using broad tolerance windows (e.g 40, 70, or 150 ms), reporting very high sensitivities such as 99.88% in LUDB and 99.48% in QTDB[27]. However, such thresholds are substantially less stringent than the temporal precision assessed in our study, as predictions with timing errors of several tens of milliseconds may still be considered correct. Consequently, these metrics can obscure clinically and methodologically meaningful differences in delineation accuracy. While these previous studies commonly used public delineation datasets such as QTDB[28] and LUDB[29], our dataset (UKB) includes a larger number of recordings, analysis across 11 independent leads, and we demonstrate associations to clinical outcome data. These features enable complementary evaluation of delineation performance across diverse ECG morphologies, while also assessing the clinical relevance of derived phenotypes.

The targeted expert review of distribution extremes provided additional insight into algorithmic behaviour. Because extreme ECG interval values, including PR, QRS, and QT durations, are clinically important and associated with adverse cardiovascular outcomes, accurate delineation at the tails of the distribution is particularly relevant. This represents an important aspect of methodological validation often omitted in previous ECG delineation studies. Following application of the trained model across the full UKB ECG dataset, we therefore performed a secondary validation focused specifically on individuals at the extremes of the interval distributions to assess real-world performance at population scale. Most outlier categories demonstrated high validity (>80%), supporting robustness across the phenotypic spectrum. The lower validity observed for extremely short QT intervals highlights a recognised challenge in delineating early T-wave termination and identifies an area for further methodological refinement. Together with the development of the expert-curated annotation dataset, these findings support the reliability of the framework for large-scale ECG phenotyping and downstream epidemiological analyses.

Although this framework was developed and calibrated specifically for the UKB 12-lead ECG dataset, the underlying deep-learning architecture is modular and adaptable. Progress in automated ECG delineation remains limited by the scarcity of large-scale annotated datasets and by heterogeneity in acquisition systems, sampling frequencies, lead configurations, and annotation standards, which often reduces cross-dataset generalisability. By constructing and validating a UKB-specific reference standard, we aimed to maximise internal validity for large-scale population phenotyping. At the same time, the computationally efficient multiscale convolutional architecture can be retrained or fine-tuned in other cohorts with minimal modification. Large-scale ECG resources are increasingly becoming available across population and clinical studies, including the China Kadoorie Biobank, the UKB Cardiac Monitoring Study, and linked electronic health record–based cardiovascular resources such as the Barts BioResource. As these datasets continue to expand, including wearable-derived ECG recordings[30], there is increasing need for transparent and validated delineation frameworks that support reproducible and harmonised ECG phenotyping across cohorts. The curated annotation dataset may also provide a foundation for future self-supervised and foundation-model approaches capable of capturing more subtle waveform morphology and cross-lead electrophysiological relationships – a possible state-of-the-art approach to ECG delineation[31,32].

The reference intervals were derived from UKB participants aged 45–74 years who were predominantly of white European ancestry and may not fully generalise to other populations. In addition, UKB participants are community volunteers and are generally healthier than hospital-based clinical cohorts, which may limit extrapolation to higher-risk clinical populations.

In summary, this study establishes a validated and openly available framework for automated ECG interval measurement in the UKB, providing a standardised foundation for reproducible phenotyping and future large-scale investigations into the biological and clinical determinants of cardiac conduction and repolarisation.

## Data Availability

The curated dataset of ECG annotations and the pre-trained DL segmentation model developed in the present work will be made available as a UKB resource. The supporting scripts for training, evaluating and deploying the model will be made available via a GitHub repository.

## Ethics statement

UKB has approval from the North West Multi-centre Research Ethics Committee (MREC) as a Research Tissue Bank (REC reference: 21/NW/0157), and all participants provided written informed consent for data collection, linkage, and use in health-related research. This study was conducted using data from UK Biobank under approved application 8256. All analyses were performed in accordance with the UK Biobank Ethics and Governance Framework and relevant institutional guidelines and regulations.

