## Supplemental Material for "A curated reference dataset and deep learning model for multi-lead electrocardiographic interval measurements in UK Biobank"

### Supplemental Methods

*Annotation Calibration and GUI Optimisation*

Prior to annotating the 125 assigned ECGs, all reviewers completed a structured calibration exercise in which they independently annotated the same 30 recordings to harmonise waveform delineation criteria and minimise inter-observer variability. This process also informed optimisation of the graphical user interface to enhance precision and efficiency, including improved multi-lead alignment, incorporation of tangent guidelines, and rapid lead switching to facilitate cross-lead comparison.

The following annotation protocol was adopted:

- P-wave identification: P-waves were preferentially marked using lead II or V1, where atrial activity is typically most clearly visualised. Reviewers ensured that the resulting PR interval was physiologically plausible (generally 120–200 ms); markings implying substantially prolonged PR intervals were re-evaluated.
- QRS onset and offset: QRS onset was defined using the lead demonstrating the clearest initial negative deflection (Q-wave) or earliest deviation from baseline. QRS offset was marked in the lead with the most distinct terminal deflection before return to baseline.
- T-wave delineation: Because T-wave onset can be difficult to define, a tangent method applied to the initial upslope was used, assuming approximate waveform symmetry. T-wave offset was also determined using the tangent method. If T-wave morphology was indistinct in a given lead, reviewers relied on alternate leads where repolarisation was more clearly visualised rather than forcing annotation in low-clarity traces. Tangent guide lines were incorporated into the interface to support consistent delineation of waveform boundaries.

Following consensus on delineation criteria and optimisation of the graphical user interface, mean absolute pairwise differences of <10 ms for PR, QRS, and T-wave boundaries were considered sufficient to proceed with formal annotation of the 125 assigned ECGs.

*Deep learning segmentation model*

In brief, the model progressively extracts features across increasing temporal scales, enhances the higher-level features, and predicts class labels per time-step (PR, QRS, QT or neither). The model architecture is visualized in Supplemental Figure 1. Specifically, the network consists of 7 convolutional blocks, the first 5 with 64 filters and a kernel size increasing from 7 to 300 to capture progressively longer (multi-scale) temporal patterns, and the final 2 blocks were dilated convolutions (rates 2 and 4) with 128 filters (15 and 75 kernel size) to focus on mid/long-range patterns and enhancing higher-level features. This is followed by a fully connected layer with dropout, before a final dense layer and Softmax activation to output the probability distribution over time steps.

The Keras implementation of Hyperband was used to confirm inclusion of the smallest and largest kernel size blocks, and the dilated convolutional blocks. It was also used to determine the level of dropout after the first dense layer (0.3) and type of activation layer (ReLU).

We used a typical batch size of 64, learning rate of $10^{-5}$, early stopping criterion of 10 epochs, reduced learning rate on 5 epochs of plateaued loss (factor of 0.5, minimum rate $10^{-7}$), and a maximum of 100 epochs. The loss function was categorical cross entropy, and we used the Adam optimizer.

Input was the median ECG beat for a single lead, scaled within individual waveforms to bring the voltage into a unit interval (standard scaling). The model output—a probability map across labels per time-step—underwent several simple stages of post-processing to derive PR, QRS complex and QT intervals. First, output probabilities were set to 0 in invalid windows depending on the label: after time step 400 for P-waves, before step 250 or after step 750 for the QRS complex, or before sample 400 for the T-wave. Second, the argmax of probabilities was taken per time-step across the 4 labels, creating a 1D label sequence. Third, a median filter with windows of 50 samples was applied to the label sequence. Fourth, onsets and offsets were extracted according to the first and final indices in the label sequence, allowing for the calculation of intervals. Note that it is possible there are no valid onsets/offsets for a given wave, possibly due to insufficient amplitude in the respective region. Finally, annotations were filtered where there was invalid monotonicity of onsets and offsets between segments (e.g. QRS onset prior to the P-wave offset). This could be caused in high entropy regions where the morphology of waves is unclear or unusual, leading to irregular label sequences. Invalid annotations were generated for 5.7% of the waveforms (N=46,036), hence filtering 98 participants with invalid annotations for all leads (just 0.13% of N=73,247 participants), leaving N=73,149 participants with at least one annotated lead.

*Wavelet-based method*

The wavelet-based segmentation algorithm[1] was implemented using custom software in MATLAB R2022b (The MathWorks Inc.) in accordance with a recent study[2]. The implementation included a post-processing thresholding step where intervals were limited to the following ranges: QRSw (50, 300), PR (80, 320) and QT (200, 600).

*Neurokit method*

Annotations were created using the ecg_process function in NeuroKit(v0.2.10)[7] an automated pipeline that performs signal pre-processing, QC, QRS complex delineation and phase determination. This function requires the ECG length to equal or exceed 4 times the sampling frequency, so the median derived beat is tiled 4 times sequentially. The inferred R peak with maximal assigned "quality" was used as the basis for extracting segment onsets and offsets – quality is a continuous index generated by interpolating QRS segment distances from an averaged segment.

**Supplemental Table 1: Diagnostic and procedural codes used to define the atrial fibrillation outcome.**

| **Atrial Fibrillation** | |
|---|---|
| *ICD10 codes* | |
| I48 | Atrial fibrillation |
| I48.0 | Paroxysmal atrial fibrillation |
| I48.1 | Persistent atrial fibrillation |
| I48.2 | Chronic atrial fibrillation |
| I48.3 | Typical atrial flutter |
| I48.4 | Atypical atrial flutter |
| I48.9 | Atrial fibrillation and atrial flutter, unspecified |
| *Operative procedures* | |
| K62.1 | Percutaneous transluminal ablation of pulmonary vein to left atrium conducting system |
| K62.2 | Percutaneous transluminal ablation of atrial wall for atrial flutter |
| K62.3 | Percutaneous transluminal ablation of conducting system of heart for atrial flutter NEC |
| K62.4 | Percutaneous transluminal internal cardioversion NEC |

**Supplemental Table 2: ICD-9/ICD-10 diagnostic codes and OPCS-4 procedural codes used to define major adverse cardiovascular outcomes (MACE).** MACE outcomes were defined as a composite endpoint including myocardial infarction, heart failure, and life-threatening ventricular arrhythmias.

| **Myocardial Infarction** | |
|---|---|
| *ICD10 codes* | |
| I21 | Acute myocardial infarction |
| I21.0 | Acute transmural myocardial infarction of anterior wall |
| I21.1 | Acute transmural myocardial infarction of inferior wall |
| I21.2 | Acute transmural myocardial infarction of other sites |
| I21.3 | Acute transmural myocardial infarction of unspecified site |
| I21.4 | Acute subendocardial myocardial infarction |
| I21.9 | Acute myocardial infarction, unspecified |
| I22 | Subsequent myocardial infarction |
| I22.0 | Subsequent myocardial infarction of anterior wall |
| I22.1 | Subsequent myocardial infarction of inferior wall |
| I22.8 | Subsequent myocardial infarction of other sites |
| I22.9 | Subsequent myocardial infarction of unspecified site |
| I23 | Certain current complications following acute myocardial infarction |
| I23.0 | Haemopericardium as current complication following acute myocardial infarction |
| I23.1 | Atrial septal defect as current complication following acute myocardial infarction |
| I23.2 | Ventricular septal defect as current complication following acute myocardial infarction |
| I23.3 | Rupture of cardiac wall without haemopericardium as current complication following acute myocardial infarction |

| | |
|---|---|
| I23.4 | Rupture of chordae tendineae as current complication following acute myocardial infarction |
| I23.5 | Rupture of papillary muscle as current complication following acute myocardial infarction |
| I23.6 | Thrombosis of atrium , auricular appendage and ventricle as current complications following acute myocardial infarction |
| I23.8 | Other current complications following acute myocardial infarction |
| *Operative procedures* | |
| K40 | Saphenous vein graft replacement of coronary artery |
| K40.1 | Saphenous vein graft replacement of one coronary artery |
| K40.2 | Saphenous vein graft replacement of two coronary arteries |
| K40.3 | Saphenous vein graft replacement of three coronary arteries |
| K40.4 | Saphenous vein graft replacement of four or more coronary arteries |
| K40.9 | Unspecified saphenous vein graft replacement of coronary artery |
| K41 | Other autograft replacement of coronary artery |
| K41.1 | Autograft replacement of one coronary artery NEC |
| K41.2 | Autograft replacement of two coronary arteries NEC |
| K41.3 | Autograft replacement of three coronary arteries NEC |
| K41.4 | Autograft replacement of four or more coronary arteries NEC |
| K42 | Allograft replacement of coronary artery |
| K42.4 | Allograft replacement of four or more coronary arteries |
| K44 | Other replacement of coronary artery |
| K44.1 | Replacement of coronary arteries using multiple methods |
| K44.2 | Revision of replacement of coronary artery |
| K44.9 | Unspecified other replacement of coronary artery |
| K45 | Connection of thoracic artery to coronary artery |
| K45.1 | Double anastomosis of mammary arteries to coronary arteries |
| K45.2 | Double anastomosis of thoracic arteries to coronary arteries NEC |
| K45.3 | Anastomosis of mammary artery to left anterior descending coronary artery |
| K45.4 | Anastomosis of mammary artery to coronary artery NEC |
| K45.5 | Anastomosis of thoracic artery to coronary artery NEC |
| K45.6 | Revision of connection of thoracic artery to coronary artery |
| K45.8 | Other specified connection of thoracic artery to coronary artery |
| K45.9 | Unspecified connection of thoracic artery to coronary artery |
| K49 | Transluminal balloon angioplasty of coronary artery |
| K49.1 | Percutaneous transluminal balloon angioplasty of one coronary artery |
| K49.2 | Percutaneous transluminal balloon angioplasty of multiple coronary arteries |
| K49.3 | Percutaneous transluminal balloon angioplasty of bypass graft of coronary artery |
| K49.4 | Percutaneous transluminal cutting balloon angioplasty of coronary artery |
| K49.8 | Other specified transluminal balloon angioplasty of coronary artery |
| K49.9 | Unspecified transluminal balloon angioplasty of coronary artery |
| K50 | Other therapeutic transluminal operations on coronary artery |
| K50.1 | Percutaenous transluminal laser coronary angioplasty |
| K50.2 | Percutaneous transluminal coronary thrombolysis using streptokinase |

| | |
|---|---|
| K50.3 | Percutaneous transluminal injection of therapeutic substance into coronary artery NEC |
| K50.4 | Percutaenous transluminal atherectomy of coronary artery |
| K50.8 | Other specified other therapeutic transluminal operations on coronary artery |
| K50.9 | Unspecified other therapeutic transluminal operations on coronary artery |
| K75 | Percutaneous transluminal balloon angioplasty and insertion of stent into coronary artery |
| K75.1 | Percutaneous transluminal balloon angioplasty and insertion of 1-2 drug-eluting stents into coronary artery |
| K75.2 | Percutaneous transluminal balloon angioplasty and insertion of 3 or more drug-eluting stents into coronary artery |
| K75.3 | Percutaneous transluminal balloon angioplasty and insertion of 1-2 stents into coronary artery |
| K75.4 | Percutaneous transluminal balloon angioplasty and insertion of 3 or more stents into coronary artery NEC |
| K75.8 | Other specified percutaenous transluminal balloon angioplasty and insertion of stent into coronary artery |
| K75.9 | Unspecified percutaenous transluminal balloon angioplasty and insetion of stent into coronary artery |
| K59.6 | Implantation of cardioverter defibrillator using three electrode leads |
| K61.7 | Implantation of biventricular cardiac pacemaker system |
| K60.7 | Implantation of intravenous biventricular cardiac pacemaker system |

**Heart Failure**

*ICD10 codes*

| | |
|---|---|
| I13.0 | Hypertensive heart and renal disease with both (congestive) heart failure |
| I13.2 | Hypertensive heart and renal disease with both (congestive) heart failure and renal failure |
| I50 | Heart failure |
| I50.0 | Congestive heart failure |
| I50.1 | Left ventricular failure |
| I50.9 | Heart failure, unspecified |
| *ICD9 codes* | |
| 4280 | Congestive heart failure |
| 4281 | Left heart failure |
| 4289 | Heart failure, unspecified |

*Operative procedures*

| | |
|---|---|
| K59.6 | Implantation of cardioverter defibrillator using three electrode leads |
| K61.7 | Implantation of biventricular cardiac pacemaker system |
| K60.7 | Implantation of intravenous biventricular cardiac pacemaker system |

**Life threatening ventricular tachycardia**

*ICD10 codes*

| | |
|---|---|
| I47.2 | Ventricular tachycardia |
| I49.0 | Ventricular fibrillation and flutter |
| I46.0 | Cardiac arrest with successful resuscitation |
| I46.1 | Sudden cardiac death, so described |
| I46.9 | Cardiac arrest, unspecified |
| *Operative procedures* | |
| I47.0 | Re-entry ventricular arrhythmia |
| K57.6 | Percutaneous transluminal ablation of ventricular wall |
| K64.1 | Percutaneous radiofrequency ablation of epicardium |
| X50.3 | Advanced cardiac pulmonary resuscitation |
| X50.4 | External ventricular defibrillation |

| **ICD Implant (included for major adverse cardiovascular event endpoint)** | |
|---|---|
| **OPCS4** | **Definition (41200)** |
| K59 | Cardioverter defibrillator introduced through vein |
| K59.1 | Implantation of cardioverter defibrillator using one electrode lead |
| K59.2 | Implantation of cardioverter defibrillator using two electrode leads |
| K59.3 | Resiting of lead of cardioverter defibrillator |
| K59.4 | Renewal of cardioverter defibrillator |
| K59.6 | Implantation of cardioverter defibrillator using three electrode leads |
| K59.8 | Other specified cardioverter defibrillator introduced through the vein |
| K59.9 | Unspecified cardioverter defibrillator introduced through the vein |
| K72 | Other cardioverter defibrillator |
| K72.1 | Implantation of subcutaenous cardioverter defibrillator |
| K72.3 | Renewal of subcutaneous cardioverter defibrillator |

**Supplemental Table 3: Diagnostic codes used to identify cardiovascular events for the exclusion of participants with prevalent cardiovascular disease at baseline.**

| Subgroup | ICD9 code | ICD10 code | Definition |
|---|---|---|---|
| Ischemic Heart Disease | 411.1 | I20 | Unstable angina |
| | | I21 | Acute myocardial infarction |
| | 410.11 | I21.0 | Acute transmural myocardial infarction of anterior wall |
| | 410.41 | I21.1 | Acute transmural myocardial infarction of inferior wall |
| | 410.81 | I21.2 | Acute transmural myocardial infarction of other sites |
| | 410.91 | I21.3 | Acute transmural myocardial infarction of unspecified site |
| | 410.71 | I21.4 | Acute subendocardial myocardial infarction |
| | 410.91 | I21.9 | Acute myocardial infarction, unspecified |
| | | I22 | Subsequent myocardial infarction |
| | 410.01/410.11 | I22.0 | Subsequent myocardial infarction of anterior wall |
| | 410.21/410.31/410.41 | I22.1 | Subsequent myocardial infarction of inferior wall |
| | 410.51/410.61/410.81 | I22.8 | Subsequent myocardial infarction of other sites |
| | 410.91 | I22.9 | Subsequent myocardial infarction of unspecified site |
| | | I24 | Other acute ischaemic heart diseases |
| | | I24.0 | Coronary thrombosis not resulting in myocardial infarction |
| | 411.89 | I24.8 | Other forms of acute ischaemic heart disease |
| | 411.89 | I24.9 | Acute ischaemic heart disease, unspecified |
| | | I25 | Chronic ischaemic heart disease |
| | 429.2 | I25.0 | Atherosclerotic cardiovascular disease, so described |
| | 414.0 | I25.1 | Atherosclerotic heart disease |
| | | I25.2 | Old myocardial infarction |
| | 414.10/414.19 | I25.3 | Aneurysm of heart |
| | | I25.4 | Coronary artery aneurysm |
| | 414.8 | I25.5 | Ischaemic cardiomyopathy |
| | 414.8 | I25.6 | Silent myocardial ischaemia |
| | | I25.8 | Other forms of chronic ischaemic heart disease |
| | 414.8/414.9 | I25.9 | Chronic ischaemic heart disease, unspecified |
| Cardiomyopathies | | I42 | Cardiomyopathy |
| | 425.4 | I42.0 | Dilated cardiomyopathy |
| | 425.11 | I42.1 | Obstructive hypertrophic cardiomyopathy |
| | 425.18 | I42.2 | Other hypertrophic cardiomyopathy |

| | | | |
|---|---|---|---|
| | 425 | I42.3 | Endomyocardial (eosinophilic) disease |
| | 425.3 | I42.4 | Endocardial fibroelastosis |
| | 425.4 | I42.5 | Other restrictive cardiomyopathy |
| | 425.5 | I42.6 | Alcoholic cardiomyopathy |
| | 425.9 | I42.7 | Cardiomyopathy due to drugs and other external agents |
| | 425.2/425.4 | I42.8 | Other cardiomyopathies |
| | 425.4/425.9 | I42.9 | Cardiomyopathy, unspecified |
| | 425.8 | I43 | Cardiomyopathy in diseases classified elsewhere |
| | | I43.0 | Cardiomyopathy in infectious and parasitic diseases classified elsewhere |
| | 425.7 | I43.1 | Cardiomyopathy in metabolic diseases |
| | 425.7 | I43.2 | Cardiomyopathy in nutritional diseases |
| | 425.8 | I43.8 | Cardiomyopathy in other diseases classified elsewhere |
| Heart Failure | | I50 | Heart failure |
| | 428.0 | I50.0 | Congestive heart failure |
| | 428.1 | I50.1 | Left ventricular failure |
| | 428.0/428.9 | I50.9 | Heart failure, unspecified |
| Atrial Arrhythmia | 427.3 | I48 | Atrial fibrillation |
| | 427.31 | I48.0 | Paroxysmal atrial fibrillation |
| | 427.31 | I48.1 | Persistent atrial fibrillation |
| | 427.31 | I48.2 | Chronic atrial fibrillation |
| | 427.32 | I48.3 | Typical atrial flutter |
| | 427.32 | I48.4 | Atypical atrial flutter |
| | | I48.9 | Atrial fibrillation and atrial flutter, unspecified |
| Ventricular Arrhythmia | 427.1 | I47.2 | Ventricular tachycardia |
| | | I49.0 | Ventricular fibrillation and flutter |
| Significant conduction disease | | I44.1 | Atrioventricular block, second degree |
| | | I44.2 | Atrioventricular block, complete |
| Arrhythmia general | 427.9 | I49.9 | Cardiac arrhythmia, unspecified |
| Cardiac Arrest | 427.5 | I46.1 | Sudden cardiac death, so described |
| | 427.5 | I46.9 | Cardiac arrest, unspecified |
| SCD | 427.5 | I46.1 | Sudden cardiac death, so described |
| TIA | | G45 | Transient cerebral ischaemic attacks and related syndromes |
| | | G45.0 | Vertebro-basilar artery syndrome |
| | | G45.3 | Amaurosis fugax |
| | | G45.8 | Other transient cerebral ischaemic attacks and related |
| | | G45.9 | Transient cerebral ischaemic attack, unspecified |
| Ischaemic Stroke | | I63 | Cerebral infarction |
| | 434.91 | I63.0 | Cerebral infarction due to thrombosis of precerebral arteries |

| | | | |
|---|---|---|---|
| | 434.91 | I63.1 | Cerebral infarction due to embolism of precerebral arteries |
| | 434.91 | I63.2 | Cerebral infarction due to unspecified occlusion or stenosis of precerebral arteries |
| | 434.01 | I63.3 | Cerebral infarction due to thrombosis of cerebral arteries |
| | 434.11 | I63.4 | Cerebral infarction due to embolism of cerebral arteries |
| | 434.91 | I63.5 | Cerebral infarction due to unspecified occlusion or stenosis of cerebral arteries |
| | | I63.8 | Other cerebral infarction |
| | 434.91 | I63.9 | Cerebral infarction, unspecified |
| Haemorrhagic stroke | | I61 | Intracerebral haemorrhage |
| | | I61.0 | Intracerebral haemorrhage in hemisphere, subcortical |
| | | I61.1 | Intracerebral haemorrhage in hemisphere, cortical |
| | | I61.2 | Intracerebral haemorrhage in hemisphere, unspecified |
| | | I61.3 | Intracerebral haemorrhage in brain stem |
| | | I61.4 | Intracerebral haemorrhage in cerebellum |
| | | I61.5 | Intracerebral haemorrhage, intraventricular |
| | | I61.6 | Intracerebral haemorrhage, multiple localised |
| | | I61.8 | Other intracerebral haemorrhage |
| | | I61.9 | Intracerebral haemorrhage, unspecified |
| Unspecified stroke | 436 | I64 | Stroke, not specified as haemorrhage or infarction |
| Aortic/Peripheral vascular disease | | I70 | Atherosclerosis |
| | | I70.0 | Atherosclerosis of aorta |
| | | I70.00 | Atherosclerosis of aorta (without gangrene) |
| | | I70.01 | Atherosclerosis of aorta (with gangrene) |
| | | I70.2 | Atherosclerosis of arteries of the extremities |
| | | I70.20 | Atherosclerosis of arteries of extremities (without gangrene) |
| | | I70.21 | Atherosclerosis of arteries of extremities (with gangrene) |
| | | I70.8 | Atherosclerosis of other arteries |
| | | I70.80 | Atherosclerosis of other arteries (without gangrene) |
| | | I73 | Other peripheral vascular diseases |
| | | I73.0 | Raynaud's syndrome |
| | | I73.1 | Thromboangiitis obliterans [Buerger] |
| | | I73.8 | Other specified peripheral vascular diseases |
| | | I73.9 | Peripheral vascular disease, unspecified |
| | | I74 | Arterial embolism and thrombosis |
| | | I74.0 | Embolism and thrombosis of abdominal aorta |

| | | |
|---|---|---|
| | I74.1 | Embolism and thrombosis of other and unspecified parts of aorta |
| | I74.2 | Embolism and thrombosis of arteries of the upper extremities |
| | I74.3 | Embolism and thrombosis of arteries of the lower extremities |
| | I74.4 | Embolism and thrombosis of arteries of extremities, unspecified |
| | I74.5 | Embolism and thrombosis of iliac artery |
| | I74.8 | Embolism and thrombosis of other arteries |
| | I74.9 | Embolism and thrombosis of unspecified artery |
| Other vascular disease | I71 | Aortic aneurysm and dissection |
| | I71.0 | Dissection of aorta [any part] |
| | I71.1 | Thoracic aortic aneurysm, ruptured |
| | I71.2 | Thoracic aortic aneurysm, without mention of rupture |
| | I71.3 | Abdominal aortic aneurysm, ruptured |
| | I71.4 | Abdominal aortic aneurysm, without mention of rupture |
| | I71.5 | Thoracoabdominal aortic aneurysm, ruptured |
| | I71.6 | Thoracoabdominal aortic aneurysm, without mention of rupture |
| | I71.8 | Aortic aneurysm of unspecified site, ruptured |
| | I71.9 | Aortic aneurysm of unspecified site, without mention of rupture |
| Hypertensive heart disease | I11 | Hypertensive heart disease |
| | I11.0 | Hypertensive heart disease with (congestive) heart failure |
| | I11.9 | Hypertensive heart disease without (congestive) heart failure |
| Non-rheumatic valvular heart disease | I34 | Nonrheumatic mitral valve disorders |
| | I34.0 | Mitral (valve) insufficiency |
| | I34.1 | Mitral (valve) prolapse |
| | I34.2 | Nonrheumatic mitral (valve) stenosis |
| | I34.8 | Other nonrheumatic mitral valve disorders |
| | I34.9 | Nonrheumatic mitral valve disorder, unspecified |
| | I35 | Nonrheumatic aortic valve disorders |
| | I35.0 | Aortic (valve) stenosis |
| | I35.1 | Aortic (valve) insufficiency |
| | I35.2 | Aortic (valve) stenosis with insufficiency |
| | I35.8 | Other aortic valve disorders |
| | I35.9 | Aortic valve disorder, unspecified |
| | I36 | Nonrheumatic tricuspid valve disorders |

| | | | |
|---|---|---|---|
| | | I36.0 | Nonrheumatic tricuspid (valve) stenosis |
| | | I36.1 | Nonrheumatic tricuspid (valve) insufficiency |
| | | I36.8 | Other nonrheumatic tricuspid valve disorders |
| | | I36.9 | Nonrheumatic tricuspid valve disorder, unspecified |
| | | I37 | Pulmonary valve disorders |
| | | I37.0 | Pulmonary valve stenosis |
| | | I37.1 | Pulmonary valve insufficiency |
| | | I37.2 | Pulmonary valve stenosis with insufficiency |
| | | I37.8 | Other pulmonary valve disorders |
| | | I37.9 | Pulmonary valve disorder, unspecified |
| Congenital heart disease | | Q20 | Congenital malformations of cardiac chambers and connexions |
| | | Q20.0 | Common arterial trunk |
| | | Q20.1 | Double outlet right ventricle |
| | | Q20.2 | Double outlet left ventricle |
| | | Q20.3 | Discordant ventriculoarterial connexion |
| | | Q20.4 | Double inlet ventricle |
| | | Q20.5 | Discordant atrioventricular connexion |
| | | Q20.6 | Isomerism of atrial appendages |
| | | Q20.8 | Other congenital malformations of cardiac chambers and connexions |
| | | Q20.9 | Congenital malformation of cardiac chambers and connexions, unspecified |
| | | Q21 | Congenital malformations of cardiac septa |
| | | Q21.0 | Ventricular septal defect |
| | | Q21.1 | Atrial septal defect |
| | | Q21.2 | Atrioventricular septal defect |
| | | Q21.3 | Tetralogy of Fallot |
| | | Q21.4 | Aortopulmonary septal defect |
| | | Q21.8 | Other congenital malformations of cardiac septa |
| | | Q21.9 | Congenital malformation of cardiac septum, unspecified |
| | | Q22 | Congenital malformations of pulmonary and tricuspid valves |
| | | Q22.1 | Congenital pulmonary valve stenosis |
| | | Q22.2 | Congenital pulmonary valve insufficiency |
| | | Q22.4 | Congenital tricuspid stenosis |
| | | Q22.5 | Ebstein's anomaly |
| | | Q22.8 | Other congenital malformations of tricuspid valve |
| | | Q22.9 | Congenital malformation of tricuspid valve, unspecified |
| | | Q23 | Congenital malformations of aortic and mitral valves |
| | | Q23.0 | Congenital stenosis of aortic valve |
| | | Q23.1 | Congenital insufficiency of aortic valve |

| | | |
|---|---|---|
| | Q23.2 | Congenital mitral stenosis |
| | Q23.3 | Congenital mitral insufficiency |
| | Q23.4 | Hypoplastic left heart syndrome |
| | Q23.8 | Other congenital malformations of aortic and mitral valves |
| | Q23.9 | Congenital malformation of aortic and mitral valves, unspecified |
| | Q24 | Other congenital malformations of heart |
| | Q24.0 | Dextrocardia |
| | Q24.1 | Levocardia |
| | Q24.3 | Pulmonary infundibular stenosis |
| | Q24.4 | Congenital subaortic stenosis |
| | Q24.5 | Malformation of coronary vessels |
| | Q24.6 | Congenital heart block |
| | Q24.8 | Other specified congenital malformations of heart |
| | Q24.9 | Congenital malformation of the heart, unspecified |
| Myocarditis | B33.2 | Viral carditis |
| | I40.0 | Acute myocarditis |
| | I40.1 | Isolated myocarditis |
| | I40.8 | Other acute myocarditis |
| | I40.9 | Acute myocarditis, unspecified |
| | I41.1 | Myocarditis in viral diseases classified elsewhere |
| | I41.2 | Myocarditis in other infectious and parasitic diseases classified elsewhere |
| | I41.8 | Myocarditis in other diseases classified elsewhere |
| | I51.4 | Myocarditis, unspecified |

**Supplementary Table 4:** Pairwise agreement analyses between lead-averaged ECG annotations (defined as the median interval across leads) and expert (manual) annotations.

| *Annotations* | | **ICC** | | **Pearson R** | | **OLS** | |
|---|---|---|---|---|---|---|---|
| *Interval* | *Method* | **ICC [95% CI]** | ***P*-value** | **r** | ***P*-value** | **Adj. $R^2$** | ***P*-value** |
| *PR* | *Deep-learning (present work)* | 0.92 [0.91-0.94] | < .001 | 0.93 | < .001 | 0.86 | < .001 |
| | Wavelet-based | 0.88 [0.85-0.90] | < .001 | 0.88 | < .001 | 0.78 | < .001 |
| | *UKB CardioSoft* | 0.89 [0.87-0.92] | .696 | 0.90 | < .001 | 0.80 | < .001 |
| | *NeuroKit* | 0.01 [-0.20-0.21] | .465 | 0.01 | .932 | -0.01 | .932 |
| *QRS* | *Deep-learning (present work)* | 0.73 [067-0.77] | < .001 | 0.76 | < .001 | 0.57 | < .001 |
| | Wavelet-based | 0.68 [0.61-0.74] | < .001 | 0.69 | < .001 | 0.47 | < .001 |
| | *UKB CardioSoft* | 0.72 [0.66-0.77] | < .001 | 0.72 | < .001 | 0.52 | < .001 |
| | *NeuroKit* | 0.04 [-0.16-0.24] | .334 | 0.05 | < .001 | -0.01 | .625 |
| *QT* | *Deep-learning (present work)* | 0.98 [0.98-0.98] | < .001 | 0.98 | < .001 | 0.96 | < .001 |
| | Wavelet-based | 0.94 [0.93-0.95] | < .001 | 0.94 | < .001 | 0.89 | < .001 |

| | | | | | | | |
|---|---|---|---|---|---|---|---|
| | *UKB CardioSoft* | 0.95 [0.93-0.96] | < .001 | 0.95 | < .001 | 0.90 | < .001 |
| | *NeuroKit* | -0.02 [-0.22-0.18] | 0.578 | -0.02 | 0.845 | -0.01 | 0.845 |

**Supplemental Table 5:** Pairwise ECG annotation agreement analyses, between lead-specific automated annotations and expert (manual) annotations. Note that UKB CardioSoft is not included as it is limited to averaged intervals across leads.

| *Annotations* | | **ICC** | | **Pearson R** | | **OLS** | |
|---|---|---|---|---|---|---|---|
| *Interval* | *Method* | **ICC [95% CI]** | ***P*-value** | **R** | ***P*-value** | **Adj. $R^2$** | ***P*-value** |
| *PR* | *Deep-learning (present work)* | 0.89 [0.88-0.90] | < .001 | 0.89 | < .001 | 0.79 | < .001 |
| | Wavelet-based | 0.80 [0.78-0.81] | < .001 | 0.80 | < .001 | 0.64 | < .001 |
| | *NeuroKit* | 0.03 [-0.02-0.08] | .093 | 0.05 | .059 | 0.00 | .059 |
| *QRS* | *Deep-learning (present work)* | 0.68 [0.66-0.70] | < .001 | 0.70 | < .001 | 0.49 | < .001 |
| | Wavelet-based | 0.47 [0.44-0.50] | < .001 | 0.49 | < .001 | 0.24 | < .001 |
| | *NeuroKit* | 0.04 [0.01-0.08] | .011 | 0.08 | < .001 | 0.01 | .625 |
| *QT* | *Deep-learning (present work)* | 0.95 [0.95-0.95] | < .001 | 0.95 | < .001 | 0.90 | < .001 |
| | Wavelet-based | 0.88 [0.87-0.89] | < .001 | 0.88 | < .001 | 0.78 | < .001 |
| | *NeuroKit* | 0.39 [0.36-0.42] | < .001 | 0.47 | < .001 | 0.22 | < .001 |

**Supplemental Table 6**: Cox proportional hazards regression models for the association of PR intervals with AF.

| **AF** | **Univariate** | | | | **Multivariate** | | |
|---|---|---|---|---|---|---|---|
| ***Interval*** | ***Method*** | **HR [95% CI]** | ***P*-value** | **Concordance** | **HR [95% CI]** | ***P*-value** | **Concordance** |
| *PR* *N=44,985* | *Deep-learning (present work)* | 1.91 [1.56-2.33] | < **.001** | 0.534 | 1.33 [1.09-1.64] | **.005** | 0.708 |
| | *Wavelet-based* | 1.76 [1.49-2.07] | < **.001** | 0.546 | 1.31 [1.10-1.55] | **.001** | 0.708 |
| | *UKB CardioSoft* | 1.78 [1.43-2.22] | < **.001** | 0.527 | 1.24 [0.99-1.55] | .056 | 0.708 |

**Supplemental Table 7:** Cox proportional hazards regression models for the association of annotated intervals with MACE.

| MACE | | Univariate | | | Multivariate | | |
|---|---|---|---|---|---|---|---|
| ***Interval*** | ***Method*** | **HR [95% CI]** | ***P*-value** | **Concordance** | **HR [95% CI]** | ***P*-value** | **Concordance** |
| *QRS* *N=46,747* | *Deep-learning (present work)* | 2.64 [2.01-3.47] | **< .001** | 0.526 | 1.60 [1.21-2.10] | **< .001** | 0.706 |
| | *Wavelet-based* | 3.02 [2.27-4.00] | **< .001** | 0.524 | 1.89 [1.42-2.51] | **< .001** | 0.707 |
| | *UKB CardioSoft* | 2.73 [2.06-3.61] | **< .001** | 0.524 | 1.69 [1.27-2.24] | **< .001** | 0.706 |
| *QTc* *N=46,742* | *Deep-learning (present work)* | 4.74 [3.48-6.44] | **< .001** | 0.525 | 2.91 [2.13-3.97] | **< .001** | 0.708 |
| | *Wavelet-based* | 1.84 [1.57-2.15] | **< .001** | 0.558 | 1.67 [1.43-1.95] | **< .001** | 0.711 |
| | *UKB CardioSoft* | 1.46 [1.21-1.77] | **< .001** | 0.528 | 1.14 [0.94-1.38] | .180 | 0.703 |

**Supplemental Figures**

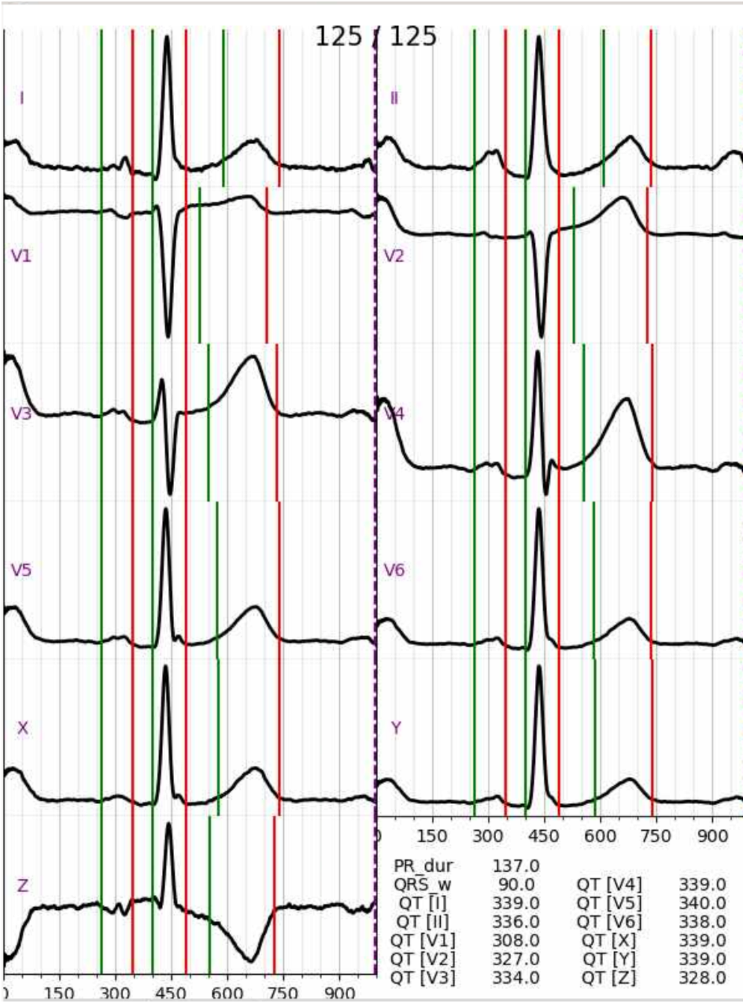


**Supplemental Figure 1:** Example of GUI. Green vertical lines indicate wave onsets and red indicate offsets. Not shown here are the tangent helper lines to determine T-wave onset and offset.

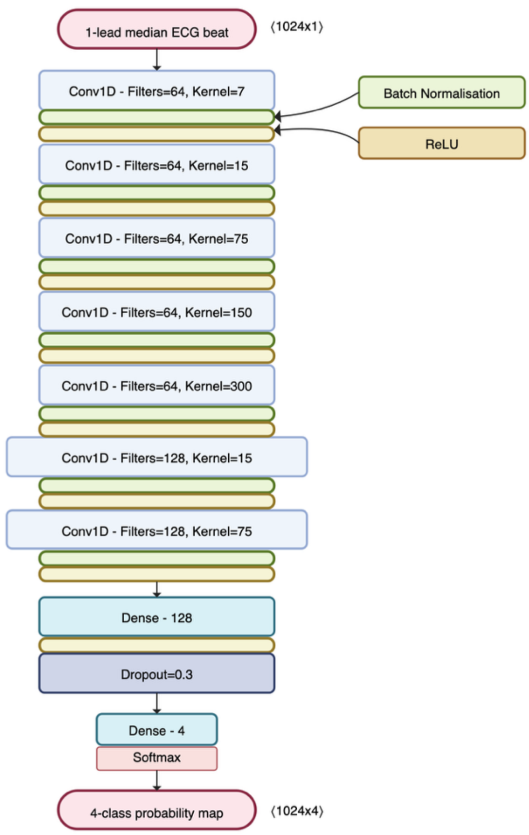


**Supplemental Figure 1:** Architecture for ECG annotation convolutional network developed in the present work. Batch normalization and ReLU activation layers are included in each 1D convolutional block and compressed for clarity. Note that the final 1D convolutions are dilated (rates 2 and 4).

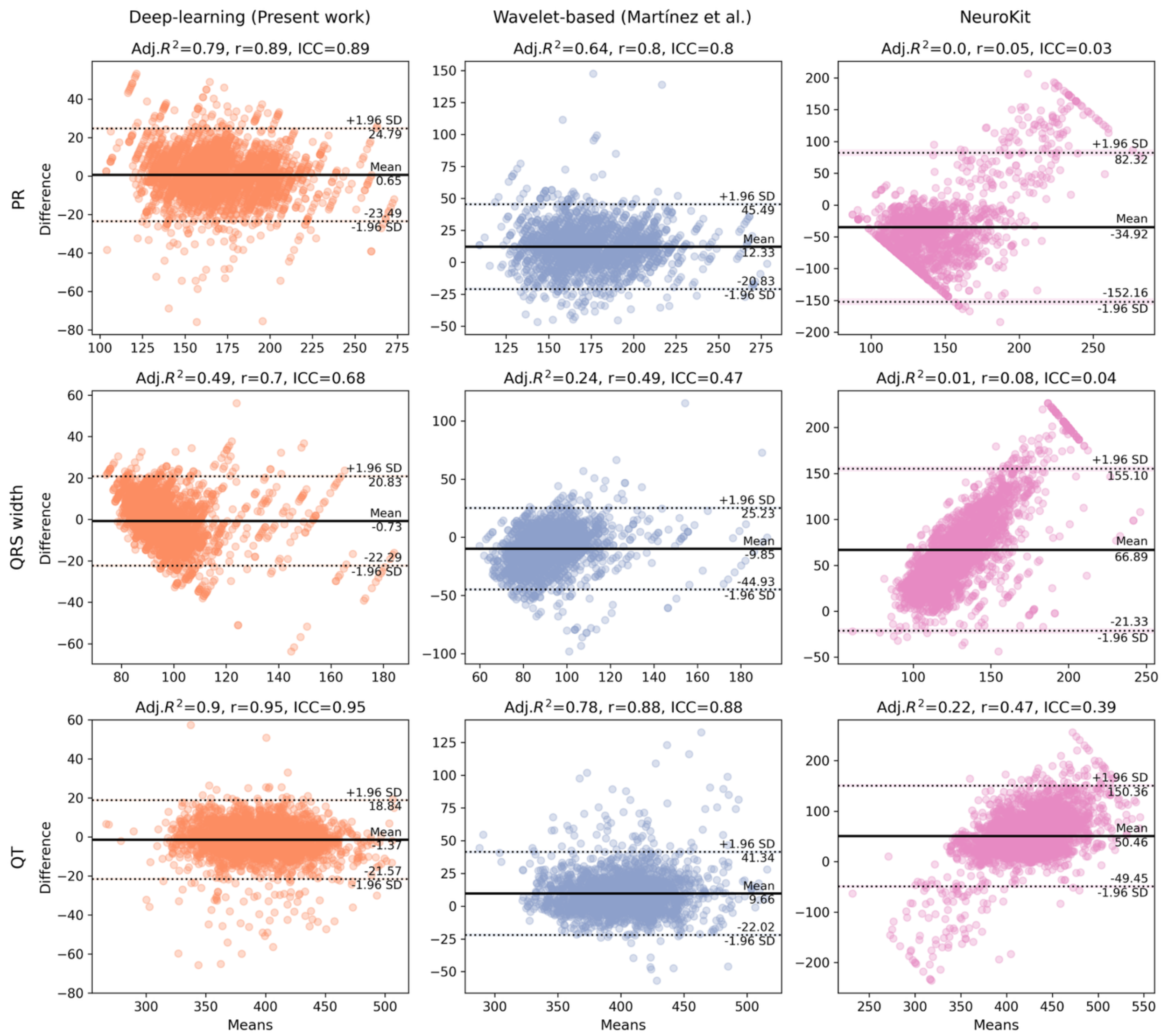


**Supplemental Figure 3:** Bland-Altman analysis for pairwise comparison of lead-specific automated and expert (manual) annotations. Corresponding agreement statistics are reported in

| *Annotations* | | ICC | | Pearson R | | OLS | |
|---|---|---|---|---|---|---|---|
| *Interval* | *Method* | **ICC [95% CI]** | ***P-value*** | **r** | ***P-value*** | **Adj. R2** | ***P-value*** |
| *PR* | *Deep-learning (present work)* | 0.92 [0.91-0.94] | < .001 | 0.93 | < .001 | 0.86 | < .001 |

| | | | | | | | |
|---|---|---|---|---|---|---|---|
| | Wavelet-based | 0.88 [0.85-0.90] | < .001 | 0.88 | < .001 | 0.78 | < .001 |
| | *UKB CardioSoft* | 0.89 [0.87-0.92] | .696 | 0.90 | < .001 | 0.80 | < .001 |
| | *NeuroKit* | 0.01 [-0.20-0.21] | .465 | 0.01 | .932 | -0.01 | .932 |
| *QRS* | *Deep-learning (present work)* | 0.73 [067-0.77] | < .001 | 0.76 | < .001 | 0.57 | < .001 |
| | Wavelet-based | 0.68 [0.61-0.74] | < .001 | 0.69 | < .001 | 0.47 | < .001 |
| | *UKB CardioSoft* | 0.72 [0.66-0.77] | < .001 | 0.72 | < .001 | 0.52 | < .001 |
| | *NeuroKit* | 0.04 [-0.16-0.24] | .334 | 0.05 | < .001 | -0.01 | .625 |
| *QT* | *Deep-learning (present work)* | 0.98 [0.98-0.98] | < .001 | 0.98 | < .001 | 0.96 | < .001 |
| | Wavelet-based | 0.94 [0.93-0.95] | < .001 | 0.94 | < .001 | 0.89 | < .001 |
| | *UKB CardioSoft* | 0.95 [0.93-0.96] | < .001 | 0.95 | < .001 | 0.90 | < .001 |
| | *NeuroKit* | -0.02 [-0.22-0.18] | 0.578 | -0.02 | 0.845 | -0.01 | 0.845 |

**Supplemental Table .**

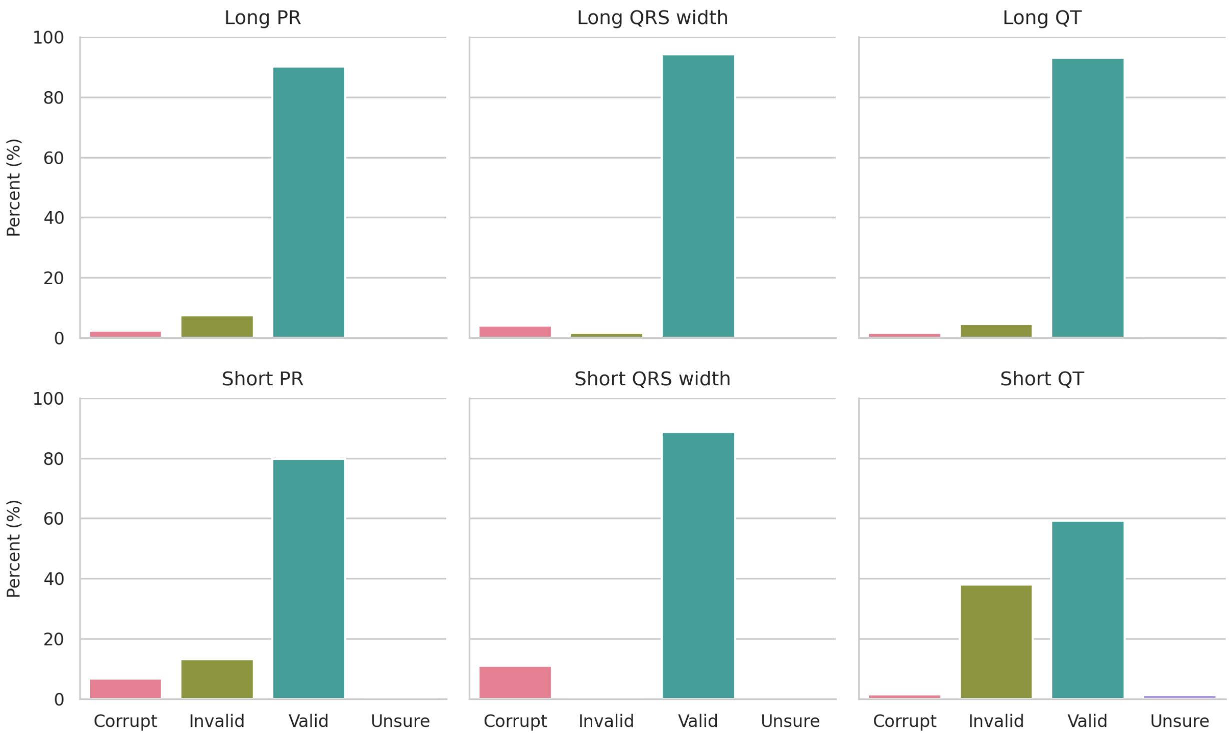


**Supplemental Figure 4:** Expert review of outlier ECG annotations, defined according to the 1% tails of annotated interval distributions generated in the present work.

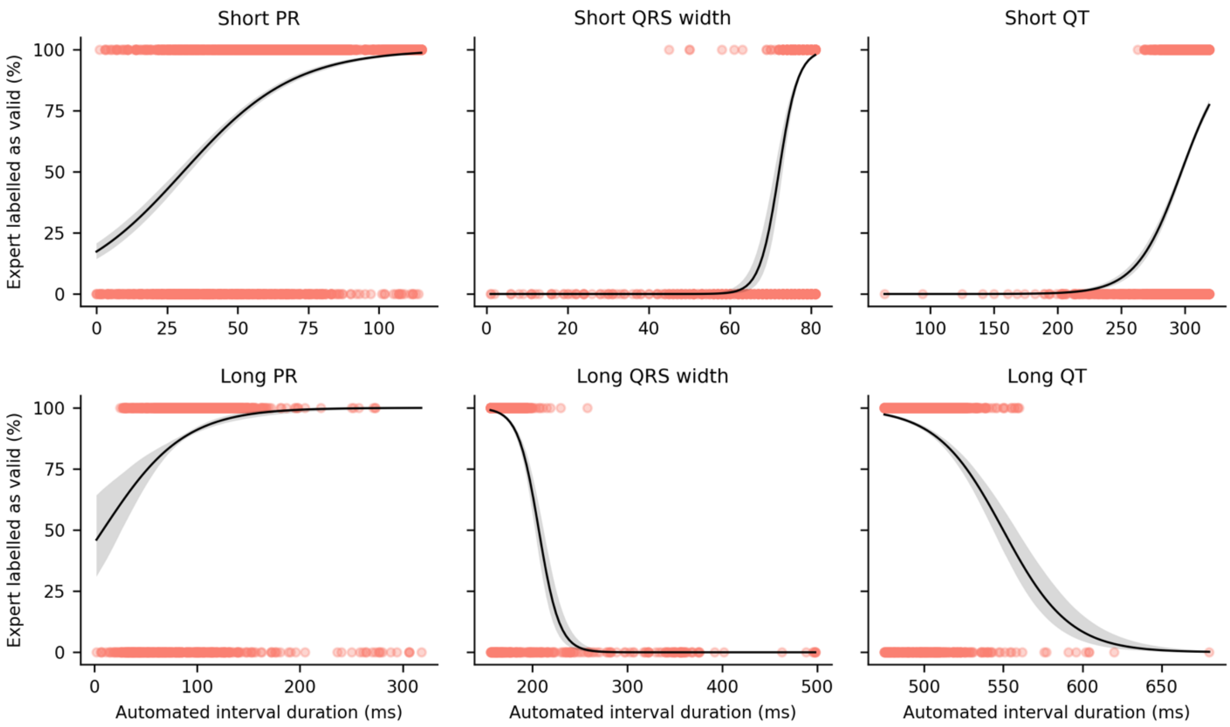


**Supplemental Figure 5:** Validity of outlier ECG annotations with respect to interval duration, with generalized linear models (binomial distribution) fit to predict expert 'valid' responses. Grey shading corresponds to 95% confidence intervals, derived from bootstrapping regression fits (N=1000).

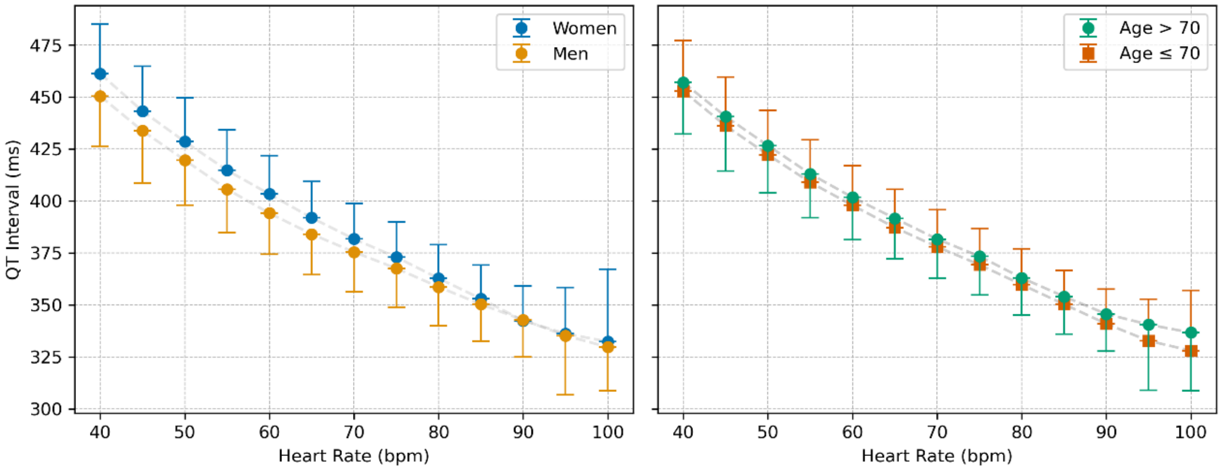


**Supplemental Figure 6:** Association between derived heart rate and automated QT interval annotations generated in the present work. Left, sex-specific annotations. Right, annotations in sub-populations with a 70 year old age cutoff. Bars correspond to a standard deviation, and dashed lines connect median intervals (dots) per heart rate bin.

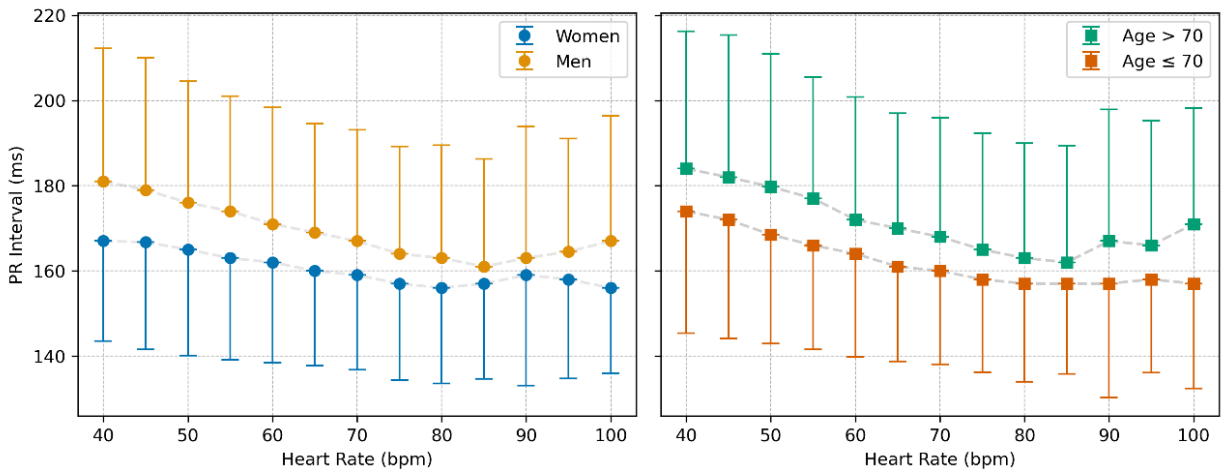


**Supplemental Figure 7:** Association between derived heart rate and automated PR interval annotations generated in the present work. Left, sex-specific annotations. Right, annotations in sub-populations with a 70 year old age cutoff. Bars correspond to a standard deviation, and dashed lines connect median intervals (dots) per heart rate bin.

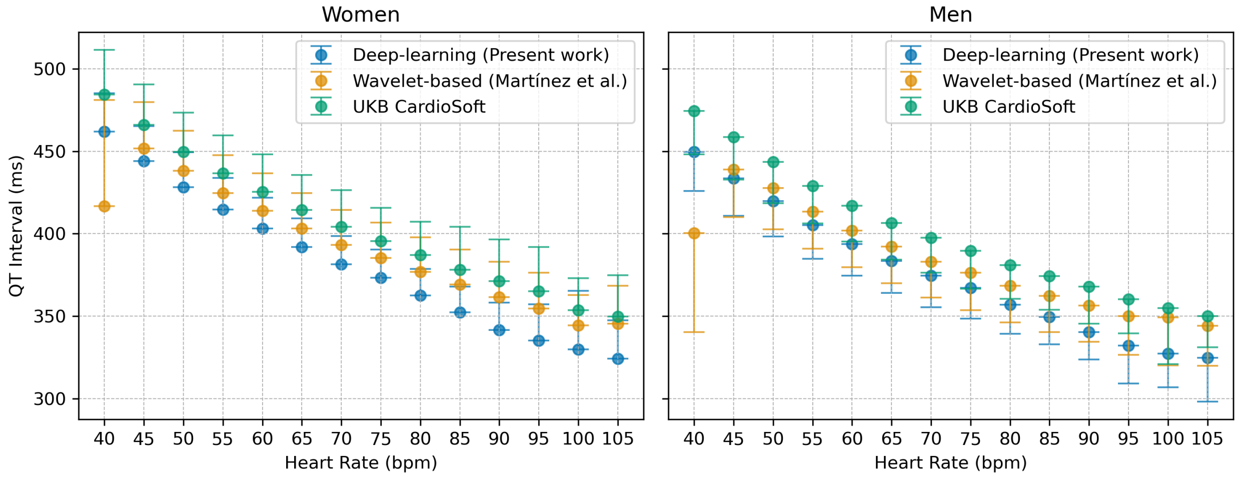


**Supplemental Figure 8:** Association between derived heart rate and automated QT interval annotations, for the most performant methods evaluated in the present study. Left, annotations for women. Right, annotations for men. Bars correspond to a standard deviation, and dashed lines connect median intervals (dots) per heart rate bin.
